# Model-based evaluation of the impact of noncompliance with public health measures on COVID-19 disease control

**DOI:** 10.1101/2020.11.29.20240440

**Authors:** Madison Stoddard, Debra Van Egeren, Kaitlyn Johnson, Smriti Rao, Josh Furgeson, Douglas E. White, Ryan P. Nolan, Natasha Hochberg, Arijit Chakravarty

## Abstract

The word ‘pandemic’ conjures dystopian images of bodies stacked in the streets and societies on the brink of collapse. Despite this frightening picture, denialism and noncompliance with public health measures are common in the historical record, for example during the 1918 Influenza pandemic or the 2015 Ebola epidemic. The unique characteristics of SARS-CoV-2—its high reproductive number (R_0_), time-limited natural immunity and considerable potential for asymptomatic spread—exacerbate the public health repercussions of noncompliance with biomedical and nonpharmaceutical interventions designed to limit disease transmission. In this work, we used game theory to explore when noncompliance confers a perceived benefit to individuals, demonstrating that noncompliance is a Nash equilibrium under a broad set of conditions. We then used epidemiological modeling to explore the impact of noncompliance on short-term disease control, demonstrating that the presence of a noncompliant subpopulation prevents suppression of disease spread. Our modeling shows that the existence of a noncompliant population can also prevent any return to normalcy over the long run. For interventions that are highly effective at preventing disease spread, however, the consequences of noncompliance are borne disproportionately by noncompliant individuals. In sum, our work demonstrates the limits of free-market approaches to compliance with disease control measures during a pandemic. The act of noncompliance with disease intervention measures creates a negative externality, rendering COVID-19 disease control ineffective in the short term and making complete suppression impossible in the long term. Our work underscores the importance of developing effective strategies for prophylaxis through public health measures aimed at complete suppression and the need to focus on compliance at a population level.

## Introduction

As we enter the unfamiliar territory of the worst global pandemic in a century, the worldwide emergence of noncompliance with public health measures aimed at limiting the spread of COVID-19 is not as surprising as it may seem at first blush (*1, 2*). During the 1918 Influenza pandemic, for example, resistance to public health measures aimed at reducing the spread of disease manifested at the individual level, leading to violence (*3*), and stiff punishments for “mask slackers” (*4, 5*). Anti-mask protesters led large demonstrations (*6*), and city councils questioned the value of mask ordinances (*7, 8*) with emotionally charged language-“under no circumstances will I be muzzled like a hydrophobic dog” (*9*). The phrasing may be dated, but the sentiment echoes precisely across a century (*10*).

For COVID-19, a number of features of the disease facilitate non-compliance. Hospitalization and death happen away from the public eye, and our changing understanding of the mechanism of transmission, the risk of mortality and the long-term consequences of the disease have favored the spread of misinformation. The spread of confusion and misinformation has been a common feature for other novel pathogen-induced pandemics such as Ebola (*1, 11, 12*) and the 1918 Flu (*13*). While the existence of pandemic denialism was easy to anticipate (*14*), the unique characteristics of SARS-CoV-2 amplify its effect. Studies suggest that asymptomatic or presymptomatic patients account for up to 40% of SARS-CoV-2 transmission (*15*), severely limiting the utility of more traditional and intuitive disease control measures such as symptomatic isolation (*16*). The high reproductive number (R_0_) of SARS-CoV-2 (reported to be 5.7 in the early days of the pandemic in Wuhan (*17*)) creates the potential for explosive growth in situations where the virus has not been completely eradicated, as has been demonstrated by a massive second wave in many European countries (*18, 19*). Making matters worse, estimates for natural immunity as a consequence of SARS-CoV-2 infection range from six to twenty-four months (*20*–*22*), creating the potential for multiple waves of disease in the short term.

Thus, the unique characteristics of SARS-CoV-2 raise the possibility that noncompliance with public health measures may create conditions that make disease control in the short term impossible or prevent any return to pre-pandemic normalcy in the long run. With this in mind, we asked three questions: First, in the specific case of SARS-CoV-2, are there circumstances that lead to a perceived benefit to noncompliance with public health measures for a substantial portion of the population? Second, what is the impact of noncompliance on the attainability of complete disease suppression for SARS-CoV-2? Third, what is the magnitude of the negative externality (a cost incurred by them that is not of their choosing) created for the compliant population as a result of noncompliance of others?

We approached the first question from the perspective of game theory, which has previously been applied to decision-making around vaccine uptake. A number of studies have examined noncompliance with measures to control COVID-19 through a social-sciences lens, exploring social and psychological risk factors associated with this behavior. These studies, from a range of different countries, have linked noncompliance to Dark Triad traits ((i.e., Machiavellianism, Psychopathy Factor 1, and narcissistic rivalry (*23*)), antisocial behaviors (*24*), higher levels of impulsivity (*25*) and a prior record of delinquent behaviors (*26*). A positive, rather than normative, framing of the question involves exploring the set of conditions for which the perceived benefit of noncompliance to the individual is simply greater than the perceived benefit of compliance. This allows us to examine the problem of compliance from the limited perspective of individuals optimizing for their own benefit without accounting for the common good, particularly relevant in the context of arguments based on personal liberty being used as a justification for noncompliance (*27*).

For the next two questions, we used a Susceptible-Exposed-Infected-Recovered-Susceptible (SEIRS) epidemiological modeling framework with a duration of immunity ranging from six to twenty-four months to explore the range of levels of compliance and intervention efficacy required for disease suppression. Our intent in this study was to establish a link between the free optimization of individuals’ outcomes as a result of noncompliance, the externalities generated by those choices, and the implications for epidemic control in the short and long term.

## Methods

### Game Theory Modeling of Compliance with COVID-19 Interventions

For the purposes of this work, we define an “intervention” as being a public health measure that reduces the transmission of COVID-19. This may be a nonpharmaceutical intervention, such as masks, or a biomedical intervention, such as a vaccine. Compliance with an intervention is defined as a binary choice. An individual can choose whether or not to comply with an intervention based on the perceived costs and benefits of the intervention. We modeled this choice using a game theoretic framework, which compares the perceived cost of compliance (reduction of quality of life resulting from the intervention) in relation to perceived cost of infection (risk-weighted morbidity/mortality burden) to the individual. Individuals derive a benefit or cost (i.e., a payoff) from interactions with other individuals in the population, who can also either be compliers or noncompliers.

We sought to determine the conditions under which noncompliance is the Nash equilibrium, or optimal behavior strategy for individuals seeking to maximize their own payoff. In a Nash equilibrium, the expected payoff to noncompliers is higher than the payoff to compliers when interacting with any other individual in the population (*28*).

For this two-strategy “game”, the payoffs to compliers and noncompliers are given in Table 1, where *q* is the cost of the intervention, *αi* is the fraction of infected individuals of type *i*, and *mi* is the perceived cost of infection for type *i* individuals, where *i* can either be *u* (noncompliers) or *v* (compliers). The cost *m*_*i*_ is the perceived risk of a negative health outcome given exposure to an infected individual. Other parameter definitions are given in Table 2. As in the SEIRS model, the efficacy of the intervention in protecting the individual from getting infected (b) is equal to the efficacy in preventing transmission (c) (i.e. *b* = *c*).

**Table 1:** Payoff matrix for compliers/noncompliers.

|  | Noncompliant interaction partner | Compliant interaction partner |
| --- | --- | --- |
| Noncomplier payoff | $-\alpha_u m_u$ | $-\alpha_v m_v c$ |
| Complier payoff | $-q - \alpha_u m_u b$ | $-q - \alpha_v m_v bc$ |

**Table 2:** Model parameters for SEIRS model.

| Parameter | Symbol | Value | Source |
| --- | --- | --- | --- |
| Latency period | $1/\alpha$ | 3 days | (29) |
| Contact period | $1/\beta$ | 1.75 days | (17) |
| Infectious period | $1/\gamma$ | 10 days | (30) |
| Natural immunity duration | $1/\delta$ | 18 months | (31) |
| Infection fatality rate | $\sigma$ | 0.68% | (32) |
| Population birth rate | $\mu$ | 1% annually | (33) |
| Population death rate | $\lambda$ | 0.9% annually | (34) |
| Fraction compliant | f | Variable |  |
| Protective efficacy | 1-b | Variable |  |
| Transmission reduction | 1-c | Variable |  |

Noncompliance is a Nash equilibrium if and only if both of the following conditions are met:

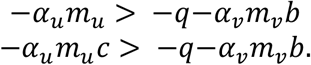

Or, equivalently

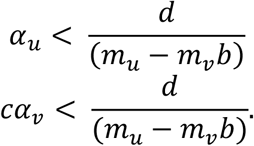

Since noncompliers are much more likely to be infected than compliers, *α*_*u*_ > *cα*_*v*_. Therefore, meeting the first condition alone (noncompliers receive a greater payoff than compliers when interacting with other noncompliers) is sufficient for noncompliance to be a Nash equilibrium.

### SEIRS Model

To support predictions of short- and long-term outcomes for the COVID-19 pandemic, we built an SEIRS (susceptible-exposed-infectious-recovered-susceptible) model to account for disease spread, waning immunity in the recovered population, and the acceptance of a vaccine or non-pharmaceutical intervention (NPI) in a fraction of the population. The model has two parallel sets of SEIR compartments representing the vaccinated or NPI-compliant (“compliant”) and unvaccinated or NPI-noncompliant (“noncompliant”) populations. The compliant population has a reduced risk of infection which is conferred by the vaccine or NPI (“protective efficacy”). The compliant population may also have a reduced risk of transmission to others upon infection resulting from physiological or behavioral changes (“transmission reduction.”) All compartments are assumed to be well-mixed, meaning that compliant and noncompliant individuals are in contact within and between groups. Vaccination or NPI compliance-based reductions in susceptibility, transmissibility, or contact rate are assumed to be time-invariant, reflecting the most optimistic case for disease control. Similarly, individuals do not move between the compliant and noncompliant compartments. Model equations (1-8) are summarized below:

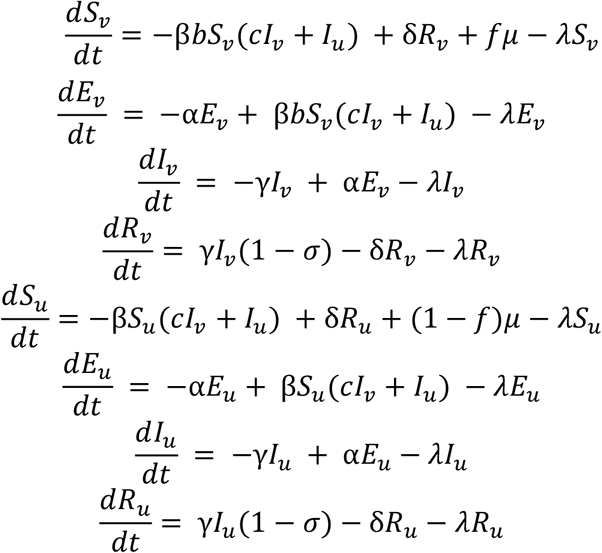

Where S represents the susceptible population, E the exposed population, I the infectious population, and R the recovered population. Subscript v represents the vaccinated or compliant sub-population, while subscript u represents the unvaccinated or noncompliant sub-population. Model parameters are summarized in Table 2.

According to the CDC, R_0_ for SARS-CoV-2 under pre-pandemic social and economic conditions is estimated to be approximately 5.7 (*17*). For the purpose of this study, an R_0_ of 5.7 is used to represent epidemiological conditions under a theoretical full return to normalcy. The contact period is derived from the relationship between the intrinsic reproductive number R_0_ and the infectious period:

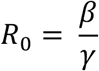

In this “normal” scenario, disease reduction interventions reduce the compliant population’s infection rate by the factor b, which represents the intervention’s protective efficacy, and the compliant population’s transmission rate by the factor c, representing the intervention’s reduction in transmissibility. For simplicity, the reduction of transmission is assumed to be equivalent to the protective efficacy (reduction of susceptibility) of each intervention. This is an optimistic assumption; in some cases, an intervention may provide little or no reduction in transmission in compliant infected individuals.

The model’s initial conditions are set to approximate current US disease prevalence and seroprevalence (as of November 2020):

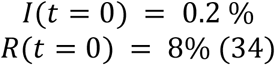

Our model lacks a seasonal component for SARS-CoV-2 transmission, as such associations have been conjectured (*35*) but not proven, and it also assumes a 18-month duration of natural immunity, as an optimistic estimate based on the duration of antibody responses seen so far months (*20*–*22*). The disease-preventing interventions and return to normalcy (which would correspond to a return to the pre-pandemic R_0_ of 5.7) are assumed to occur at the beginning of the simulation interval.

### Compliance Sweeps

To gauge the impact of NPI or vaccine compliance on population outcomes, we varied the compliant fraction under a series of simulated vaccine or NPI deployment schemes with varying degrees of protective efficacy. The model allows tracking of outcomes for the population as a whole and for the compliant and noncompliant sub-populations.

## Results

### Structural incentives for noncompliance with interventions aimed at controlling COVID-19

In Figure 1, we modeled the decision to comply with public health measures in terms of its perceived short-term impact to individuals. In game theory, a Nash equilibrium is a strategy which has a higher payoff for the individual than all other possible strategies (“no regrets”) (*28*). Individuals using a strategy that is a Nash equilibrium are unable to improve their outcome by switching strategies. Strikingly, for a large region of parameter space in this model, noncompliance is a Nash equilibrium. Even so, one can make the case that, using realistic estimates for risk of infection and risk of adverse outcomes given infection, compliance would still be a rational choice for the vast majority of the population. For example, for an intervention that is 50% effective at reducing the risk of infection, when 2% of individuals are infected, compliance is a Nash equilibrium at a 1% relative cost (ratio of the loss of quality of life associated with the intervention over the cost of infection in terms of risk of mortality, morbidity, and disability). While the decision to comply is determined by the perceived cost of infection and the perceived cost of intervention, the actual costs may be very different. The cost of wearing a mask, for example, is likely to be much less than the risk-weighted cost of death or disability due to COVID-19 (see Table S1, S2 for a more detailed analysis).

**Figure 1:**
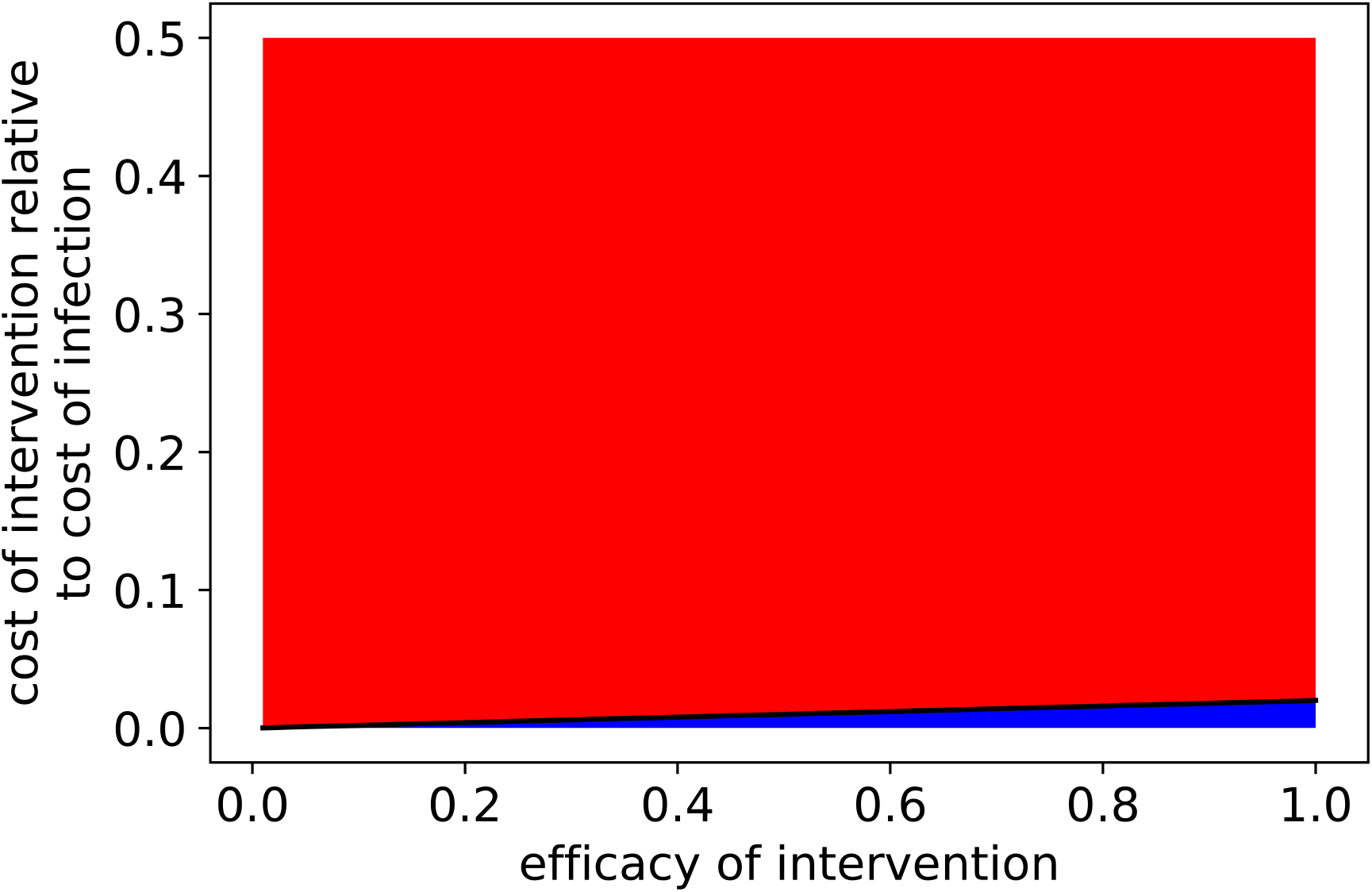
Noncompliance is a Nash equilibrium when few people are infected or when prophylaxis/therapy is more costly or less effective. Intervention efficacy and intervention cost conditions for which noncompliance is a Nash equilibrium (red) or not a Nash equilibrium (blue) if the disease is present in 2% of individuals in the population. Intervention cost relative to infection cost is defined as the ratio of intervention cost to risk-weighted infection cost

### Failure to suppress COVID-19 results in waves of disease

As shown in Figure 2, insufficient reduction in COVID-19 transmission allows the disease to persist upon a rapid return to pre-pandemic levels of social and economic activity, spreading in multiple waves over time. The model does not account for changes in behavior or environmental factors over time, so these oscillations in transmission are caused by a predator-prey dynamic within the SEIRS system rather than triggered by external factors. This dynamic is driven by the time-variant availability of susceptible hosts as immunity wanes in former COVID-19 infected individuals. Panels 2a-c represent a high compliance (95%) scenario with a 50% effective intervention. The efficacy of an intervention describes the fraction of possible transmission events it prevents. In this case, the oscillations and variability in risk for the compliant population are relatively small because the intervention serves to dampen the oscillations in transmission rate. However, in panels 2d-f, representing a low compliance (50%) scenario with a 50% effective intervention, the oscillatory pattern is much more pronounced and risk to the compliant population is variable over time (Fig. 2f). Additionally, the cumulative risk to the compliant population relative to the noncompliant population is higher when more of the population is noncompliant (Fig. 2c, f).

**Figure 2:**
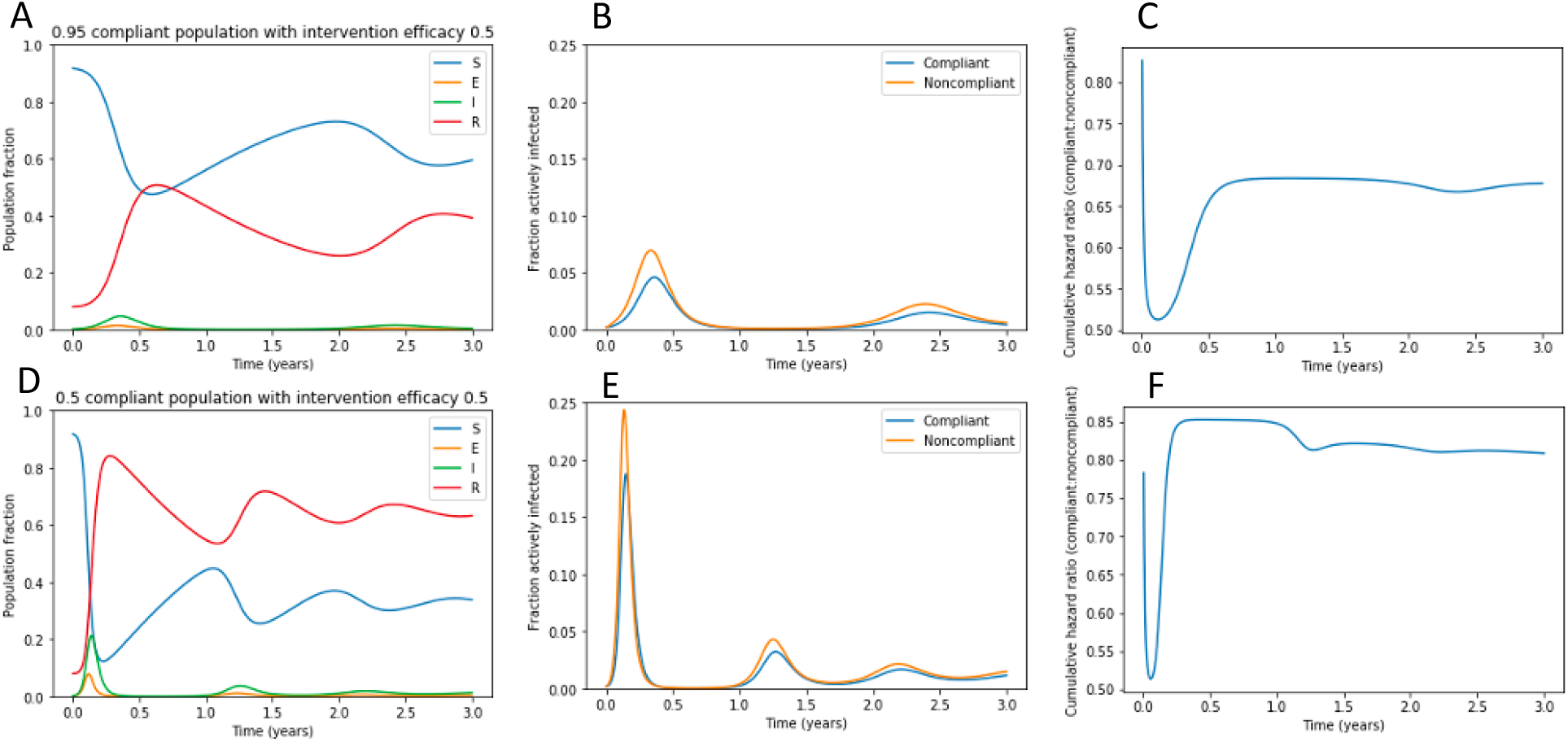
Failure to eradicate COVID-19 results in waves of disease upon a rapid return to pre-pandemic activity. Panels A and D represent the fraction of the population, including both compliant and noncompliant individuals, that is susceptible, exposed, infectious, and recovered populations over time after a return to pre-pandemic conditions under (A-C) 95% compliance or (D-F) 50% compliance with a 50% effective intervention. Panels B and E demonstrate the fraction of compliant and noncompliant individuals who are infected over time. Panels C and F demonstrate the cumulative hazard ratio for infection in noncompliant versus compliant individuals.

### Near-term suppression of COVID-19 requires a high degree of compliance

In the short term, to suppress COVID-19 while returning to pre-pandemic social and economic activity, an intervention with a high degree of efficacy and compliance is required (Fig. 3). Although effective suppression can be achieved with an intervention with as low as 65% efficacy, at least 80% compliance is required for even the most effective interventions. The predicted number of cases in the next year span three orders of magnitude, from less than one million cases to hundreds of millions of cases, depending on the effectiveness and the degree of compliance with transmission reduction interventions.

**Figure 3:**
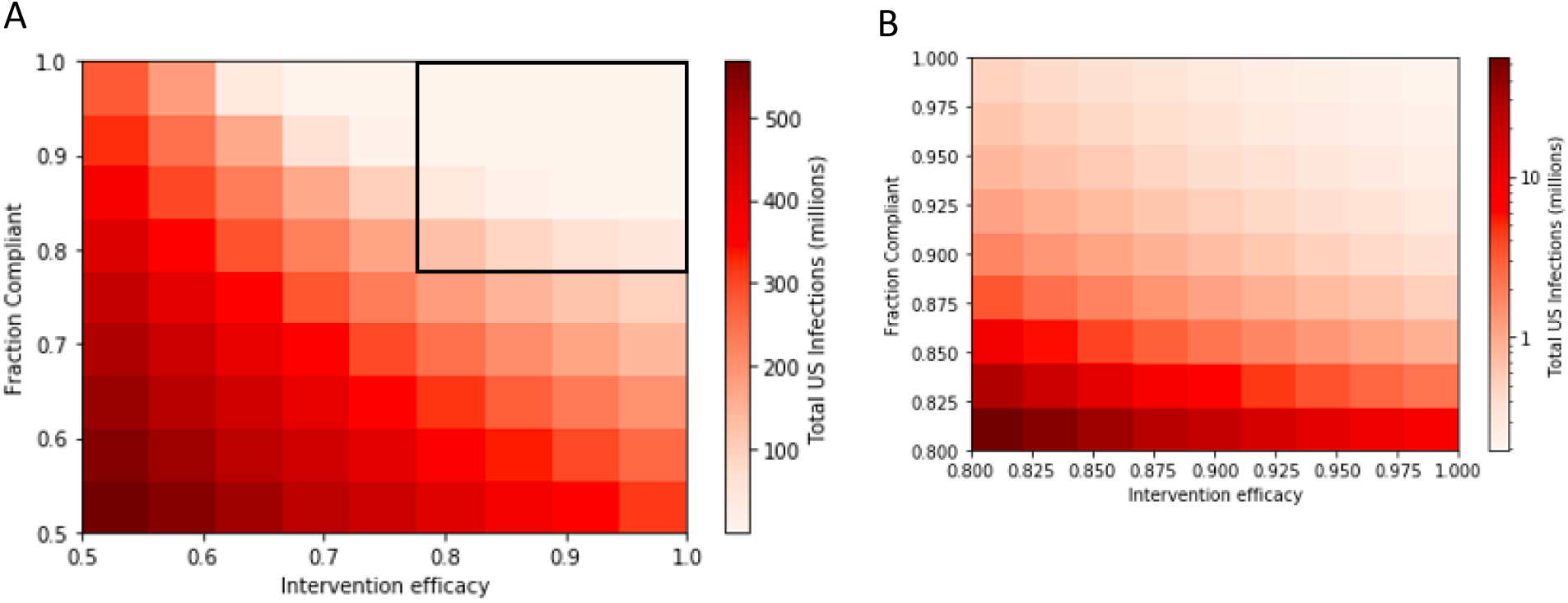
Short-term suppression of COVID-19 requires a high degree of compliance with a highly effective measure. Total US COVID-19 infections in the next year under interventions with varying efficacy and compliance. Black box on panel A shows region expanded in panel B. Panel B is displayed on a log scale.

### If COVID-19 becomes endemic, steady-state yearly spread depends on population compliance

If immunity to SARS-CoV-2 by natural infection is not life-long, as suggested by many studies (*20*–*22*), and if effective interventions are not undertaken at a large scale, the virus will become endemic. As shown in Figure 4, this means that in the long term, SARS-CoV-2 will reach a steady-state prevalence in the population. For a 50% effective intervention, the disease will become endemic even if the entire population complies with the intervention. As expected, the benefit of compliance for an individual is smaller for a 50% effective intervention (Fig. 4a) relative to a 95% effective intervention (Fig. 4b). The full compliance scenario for the 95% effective intervention is an example of disease suppression.

**Figure 4:**
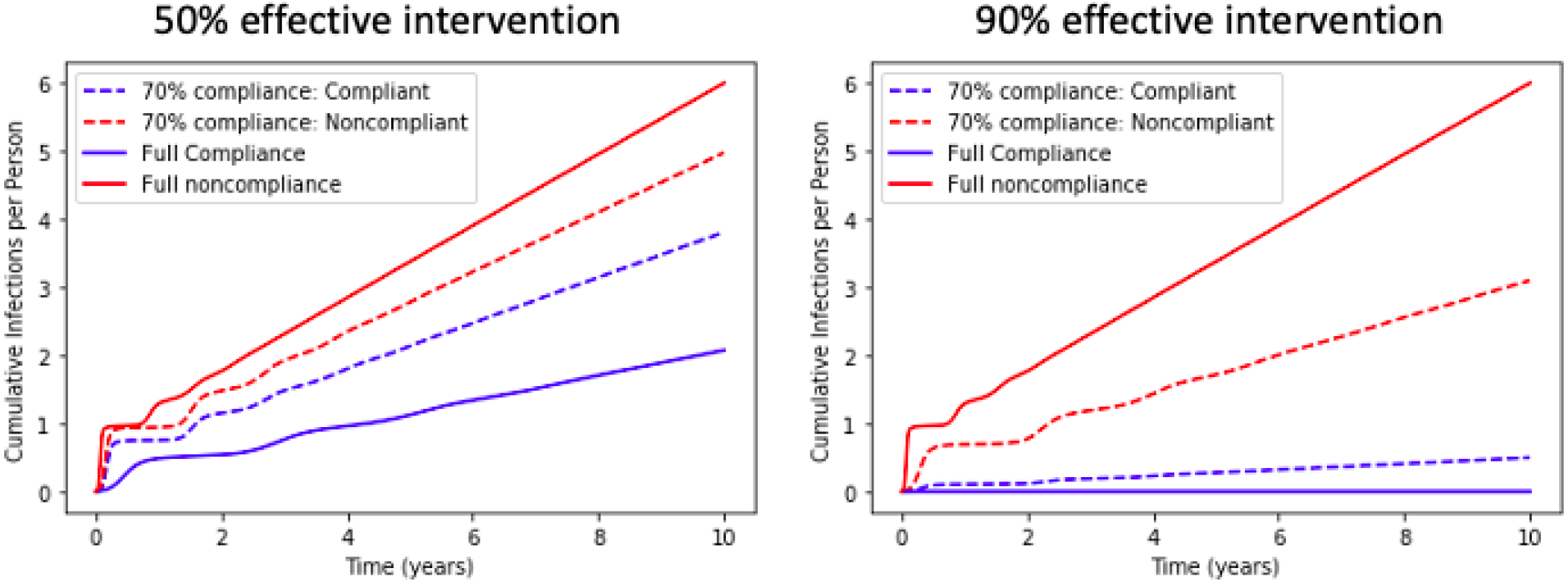
Steady-state individual risk is impacted by individual and population compliance. Cumulative infections per individual under a 50% or 90% effective intervention. Three scenarios are simulated: full noncompliance, full compliance, and 70% compliance (with outcomes for compliant and noncompliant individuals shown).

### Failure to suppress COVID-19 in the long-term results in persistent high disease burden

If complete suppression of disease is not achieved, a high annual disease burden persists indefinitely in most scenarios (Fig. 5). The marginal cost in terms of yearly cases for failures to suppress disease is highest for near-success cases and is steeply dependent on the degree of compliance (Fig. S1). This suggests that the best strategic objective for a stable return to pre-pandemic activity is complete suppression of SARS-CoV-2 spread.

**Figure 5:**
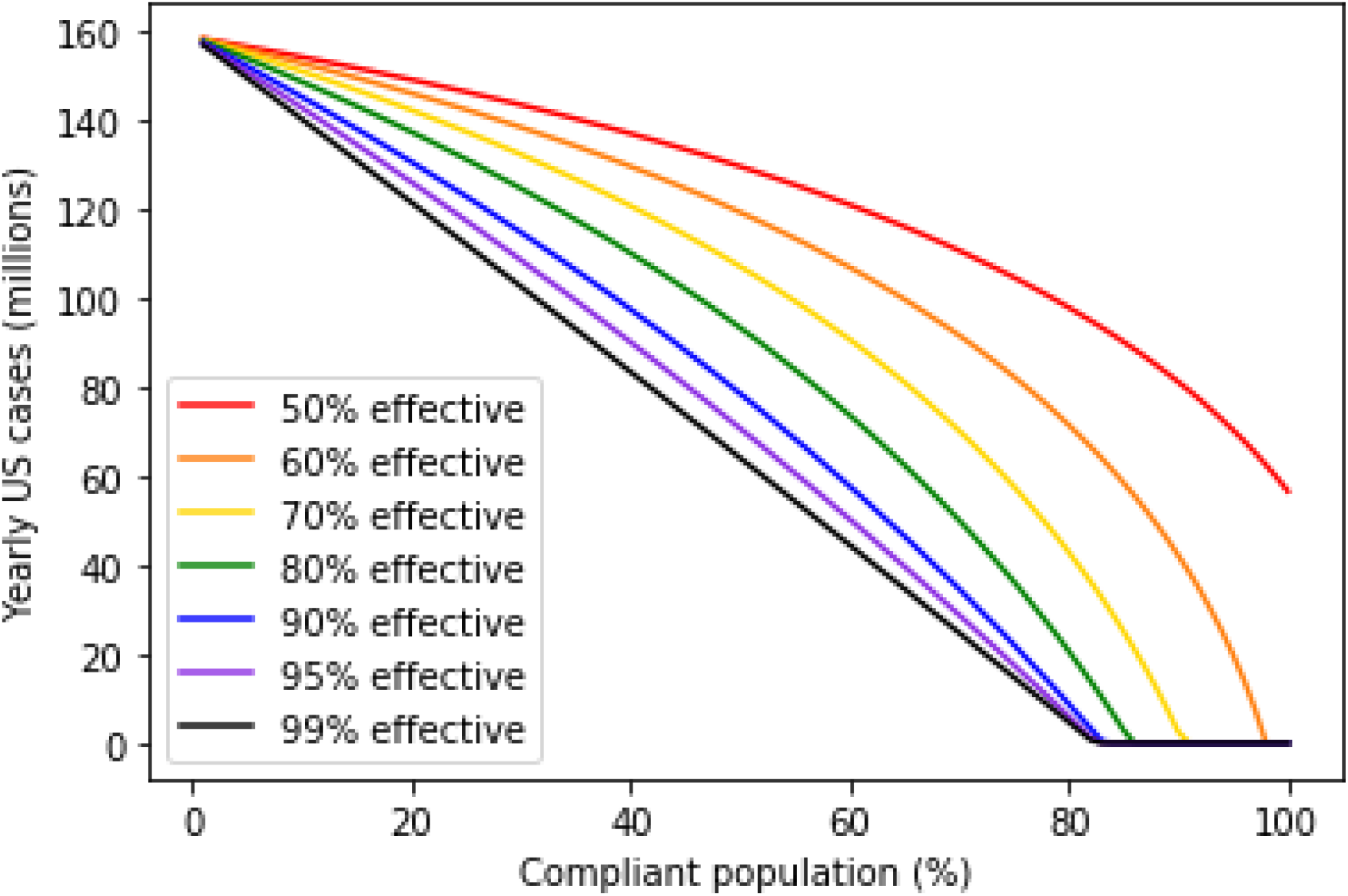
Population-level impact of interventions is highly dependent on compliance. Yearly US cases at steady-state under interventions with varying degrees of efficacy and compliance.

### Complete suppression requires high compliance and at least 60% efficacy

In Figure 5, the steady-state yearly caseload of SARS-CoV-2 is plotted against the fraction of the population complying with a variety of theoretical interventions. For interventions with greater than 60% efficacy, complete suppression can be achieved if compliance is above a certain high threshold, depending on the intervention’s efficacy. For example, with an intervention with 70% efficacy, complete suppression can be achieved with at least 92% compliance. Increasingly effective interventions reduce the compliance threshold for complete suppression and reduce the yearly caseload in endemic scenarios. However, the impact of progressive improvements in efficacy shrinks as 100% efficacy is approached. Even for a 99% effective vaccine, greater than 80% compliance is required to achieve complete suppression of SARS-CoV-2. This suggests that a high degree of intervention efficacy cannot compensate for the epidemiological impact of a large noncompliant population.

Additionally, we note that the duration of immunity does not impact these compliance thresholds for achieving complete suppression (Fig. S3). However, the duration of immunity does impact the expected yearly disease burden. To further demonstrate this point, Figures 2, 3, 4, 5, and S2 are reimplemented in the supplementary materials assuming a shorter (6-month) or longer (36-month) duration of immunity (Figs. S5-S14).

### If complete suppression is not achieved, compliant populations remain at risk without a highly effective vaccine

Although improvements in vaccine or intervention efficacy face diminishing returns on the population level and cannot fully compensate for poor compliance, compliant individuals stand to gain from even small improvements in efficacy (Fig. 6). This means that although the compliance threshold required for complete suppression may not shift substantially as efficacy improves, the incentive for individuals to comply or seek vaccination on a voluntary basis will increase as the efficacy increases.

**Figure 6:**
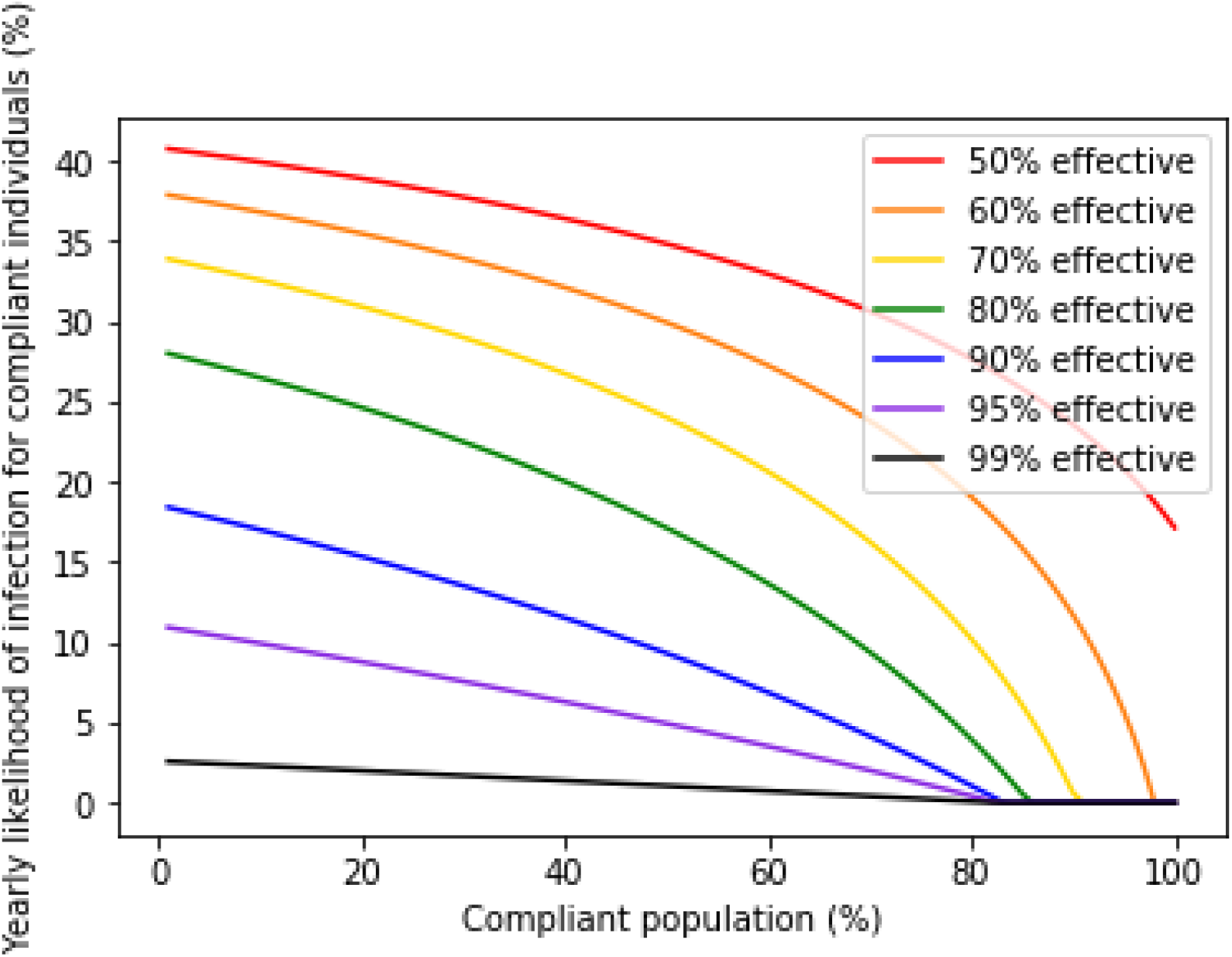
More effective interventions provide more benefit to compliant individuals if disease spread persists. Reduction in yearly likelihood of infection for compliant individuals as a function of the overall fraction of the population in compliance and the efficacy of the intervention.

## Discussion

Our work makes the case that noncompliance is embedded deeply in human nature, as individuals optimizing their own self-interest can justify their actions in terms of their own perceived cost-benefit.

Individuals may perceive noncompliance as favorable for a number of reasons (*36, 37*). They may view their own risk of being infected as lower than average (the optimism bias (*38*), which has been documented as a risk factor in predicting noncompliance for COVID-19 mitigation measures (*39*)), or they may view their own risk of adverse outcomes as a result of infection as being lower than average (*26*). Globally, the public health messaging around noncompliance has focused on the low risk of death for younger individuals (*40*–*42*) and has invoked the imagery of “shielding” highly vulnerable populations from the disease (*43*) as an altruistic motive (*44*). To the extent that many Western countries at present are facing uncontrolled disease spread, it is likely that invoking altruism may not be the most effective means of disease control. Underestimating the risk of infection may also lead to individuals believing that noncompliance is the better choice.

The interplay between risk perception and compliance is complex, and fear may also play a paradoxical role in noncompliance. A number of studies have demonstrated a link between emotions and cognitive assessment of risk. In particular, high levels of fear coupled with a low sense of efficacy may create a defensive response in individuals who then proceed to dismiss the risk (“we’re all going to die anyway”)(*45*). Studies have also shown that psychological affect plays a role in risk perception in individuals who are less comfortable and/or experienced interpreting probability (*46, 47*).

Regardless of the underlying causes, a Nash equilibrium of noncompliance creates a Tragedy of the Commons situation, where individuals acting according to their own self-interest create outcomes that are suboptimal for the common good, by spoiling the shared resource through their collective actions. The term Tragedy of the Commons dates back to an influential article written over fifty years ago (*48*), which in turn was inspired by a nineteenth-century essay describing grazing practices of farmers. Tragedy of the Commons situations are indeed common in the fields of economics, politics, environmental policy and sociology. What makes Tragedy of the Commons situations particularly intractable is that it usually only takes a small proportion of individuals optimizing for their own self-interest to create devastating externalities for the rest of the population. This behavior underscores the limitations of the laissez-faire, individualistic, approach to disease control during a pandemic. While the underpinnings of such a laissez-faire approach are often said to lie in the economic theory of utilitarianism (*49*), put forward by John Stuart Mill, such an approach actually violates the standard originally laid out by Mill by which a person’s liberty may be restricted: “The only purpose for which power can rightfully be exercised over any member of a civilized community, against his will, is to prevent harm to others” (*50*).

Given the ubiquity of the problem, some public policy solutions can be found that have close analogies to successful interventions in other spheres of human activity. First, public health messaging that seeks to alter the Nash equilibrium at an individual level are worth exploring. In individualistic societies, this may be accomplished by de-emphasizing altruism and focusing on the individual cost-benefit. One way this may be achieved is by emphasizing the long-term morbidity costs (such as cryptic heart, lung, brain and kidney damage) as have been documented to occur in even asymptomatic COVID-19 patients in an age-independent manner(*51*–*53*). An additional approach is to provide an accurate and current picture of the risk of contracting the disease. Second, public health policy that creates costs for noncompliers may serve to shift some of the externalities back on to the originator (as was the case with mask ordinances during the 1918 Flu (*4*) and fines imposed for noncompliance with COVID-19 prevention measures in some countries (*54, 55*)). Third, public health interventions should engage at the level of the community. Public health and communications experts could test a number of different messages that underscore the downside of negative externality-creating behavior at a societal level. Some of these approaches have been used previously in the context of vaccine acceptance (*56*). It is worth noting that our analysis points out a potential mechanism for the high levels of compliance observed in countries such as Korea (*57*), with strong societal norms and a positive view of COVID-19 restrictions such as mask-wearing (*58, 59*). In these cultures, the prevailing cultural beliefs may serve to lower the cost of the intervention. In this context, we note that there is a modest association (Fig. S4) between societies with strong societal norms (“tight cultures” (*60*)) and the total case count per million at this point in the pandemic (p=0.04).

From the perspective of biomedical interventions, our work points out that interventions with a high degree of protective efficacy are required for complete suppression of SARS-CoV-2, making this disease particularly challenging to control. Highly effective interventions have the dual effect of making the creation of negative externalities less beneficial for the noncompliant population (Fig. 4), and also increasing the benefit to the compliant population (Fig. 6). Notably, highly effective interventions also provide more wiggle room for public policy, as the threshold level of compliance required for the complete suppression of COVID-19 drops from approximately 95% for a minimally effective intervention to approximately 80% for highly effective interventions. Another path to disease suppression lies in implementing passive interventions that reduce the R_0_, such as improving ventilation. Such passive interventions, being not subject to the problem of individual noncompliance, can serve to lower the bar for compliance for any given intervention to achieve complete suppression (Fig. S2).

Notably, our work optimistically assumes that the impact of biomedical and nonpharmaceutical interventions is not variable over time. In some settings, “pandemic fatigue” may drive increased noncompliance with nonpharmaceutical interventions over time (*61*), and relaxations of individual caution and public health guidelines may follow improvements in regional transmission, creating reactive variability in intervention effectiveness. Although biomedical interventions such as vaccines are less susceptible to variability in day-to-day decision-making, immunity to SARS-CoV-2 is expected to wane over a period of months (*20*–*22*), which can be expected to impact the duration of vaccine protection. Challenges in vaccine distribution and compliance may compound this waning immunity, reducing the apparent effectiveness of vaccines at the individual and population scale. Additionally, we assumed that compliant individuals have a reduced risk of transmission upon infection, equal to their reduction in risk of infection. This indirect benefit is challenging to measure in clinical trials, and preclinical studies show that some vaccine candidates are capable of reducing nasal viral load (and by implication, risk of transmission) in vaccinated animals (*62, 63*), while others are not (*64, 65*).

Taken together, our work demonstrates that noncompliance with measures to control COVID-19 is at once easy to justify on an individual level and leads to devastating public health consequences. Even under optimistic assumptions about the transmission benefit and durability of preventive interventions, noncompliance presents a significant obstacle to COVID-19 suppression. Three key messages are worth keeping in mind. First, the importance of focusing on complete suppression as a desirable end goal for SARS-CoV-2 (Fig. 5) and as a prerequisite for a return to a pre-pandemic lifestyle. SARS-CoV-2 is highly transmissible and can be expected to circulate at high rates if it becomes endemic. Second, the need for public policy to focus on compliance as a key prerequisite for both short-term suppression (Fig. 3) and long-term complete suppression (Fig. 5) of COVID-19, and to seek ways to alter the space where compliance is the Nash equilibrium by increasing the cost of noncompliance. Finally, the need to focus on highly effective interventions from a biomedical perspective and to view partially effective prophylactics as contributors to the solution rather than the solution in its entirety.

It is our hope that this work draws the attention of the biomedical community to how high the bar is actually set for us to return to normalcy, and to public policymakers to highlight the need for concerted action that is focused on complete disease suppression.

## Data Availability

No patient-specific data was used, all data is available in the public domain.

