## Supplemental Materials for "Model-based evaluation of the impact of noncompliance with public health measures on COVID-19 disease control"

**Table S1.** COVID-19 Death Risk Assessment for South Dakota in August 2020. The risk of infection for a 25-year-old resulting from a decision to not wear a mask during interactions with 100 people is equivalent to driving for about one month (868 miles) while intoxicated.

| | | | |
|---|---|---|---|
| (a) | active cases | 1,358 | active cases as of 8/16/2020 (*1*) |
| (b) | population | 903,027 | |
| (c) | % population confirmed cases | 0.15% | (a) / (b) |
| (d) | % population infected | 1.09% | (c) x 7.22 (ratio of infections to cases) (*2*) |
| (e) | # interactions | 100 | user-defined parameter |
| (f) | number of expected exposures | 1 | calculated from (d) & (e) |
| (g) | p(infection \| exposure) | 4.70% | for low-risk contacts (*3*) |
| (h) | Mask reduction in risk | 85% | aOR = 0.15 (*4*) |
| (i) | p(death\| infection) for a 25-year-old | 0.004% | IFR (*5*) |
| (j) | p(death\| infection) for a 75-year-old | 8.50% | IFR (*5*) |

| | **Risk-weighted cost of infection (mortality risk)** | **Risk-equivalent drunk driving distance (mi)** |
|---|---|---|
| **For a 25-year-old** | 0.0002% | 868 (*6*) |
| **For a 75-year-old** | 0.37% | 1,844,966 (*6*) |

**Table S2.** COVID-19 Death Risk Assessment for South Dakota in November 2020. The risk of infection for a 25-year-old resulting from a decision to not wear a mask during interactions with 100 people is equivalent to driving about one year (12,000 miles) while intoxicated.

| | | | |
|---|---|---|---|
| (a) | active cases | 19,240 | active cases as of 8/16/2020 (*1*) |
| (b) | population | 903,027 | |
| (c) | % population confirmed cases | 2.13% | (a) / (b) |
| (d) | % population infected | 15.50% | (c) x 7.22 (ratio of infections to cases) (*2*) |
| (e) | # interactions | 100 | user-defined parameter |
| (f) | number of expected exposures | 15 | calculated from (d) & (e) |
| (g) | p(infection \| exposure) | 4.70% | for low-risk contacts (*3*) |
| (h) | Mask reduction in risk | 85% | aOR = 0.15 |
| (i) | p(death\| infection) for a 25-year-old | 0.004% | IFR (*5*) |
| (j) | p(death\| infection) for a 75-year-old | 8.50% | IFR (*5*) |

| | **Risk-weighted cost of infection (mortality risk)** | **Risk-equivalent drunk driving distance (mi)** |
|---|---|---|
| **For a 25-year-old** | 0.0025% | 12,301 (*6*) |
| **For a 75-year-old** | 5.26% | 26,139,277 (*6*) |

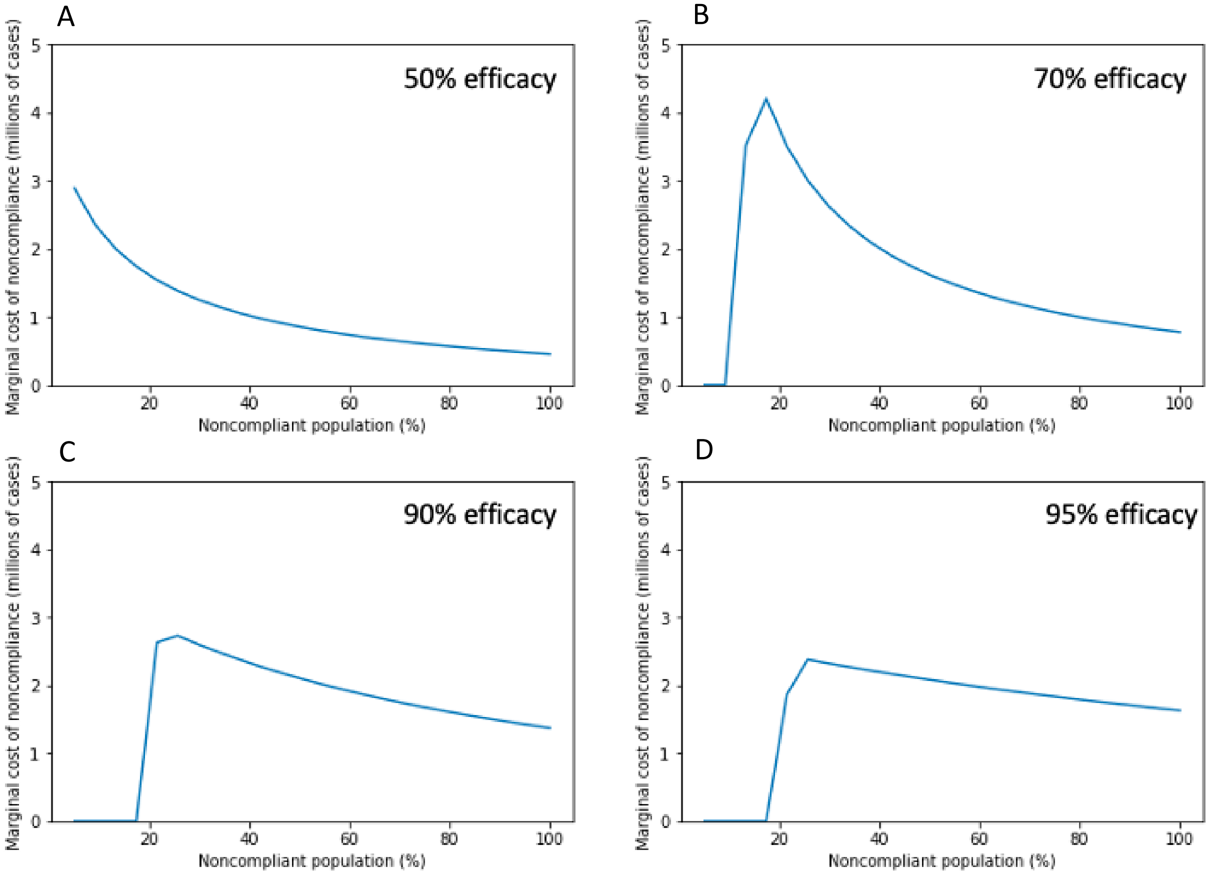


**Figure S1:** The marginal cost of noncompliance is highest as complete suppression is approached. Millions of additional yearly US cases per 1% reduction in compliance, as a function of the fraction of noncompliant individuals in the population for an intervention that is (A) 50% effective, (B) 70% effective, (C) 90% effective, or (D) 95% effective.

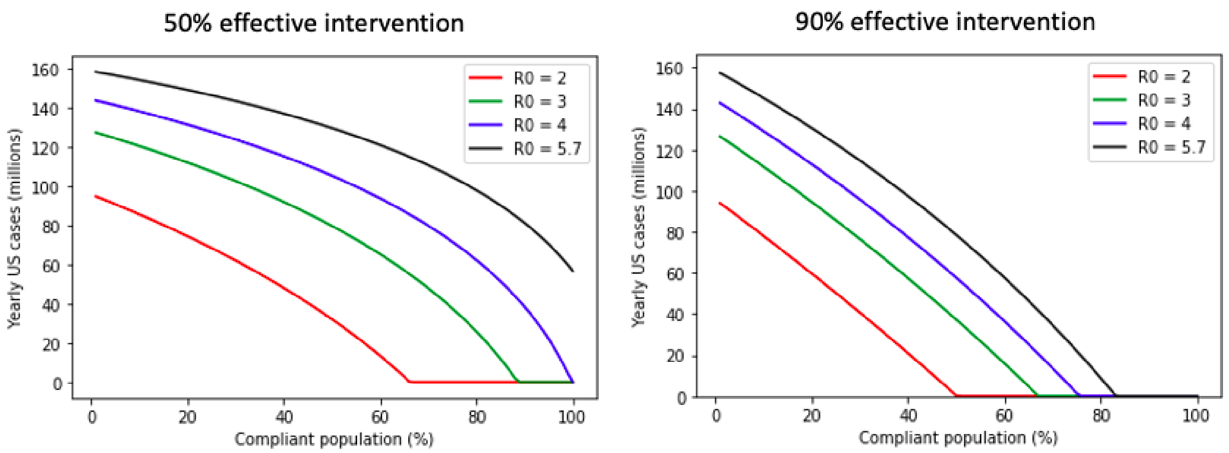


**Figure S2:** Attainability of complete suppression depends on $R_0$ and effectiveness of the intervention. Yearly US cases as a function of compliance for hypothetical scenarios in which $R_0$ = 2, 3, 4 or 5.7.

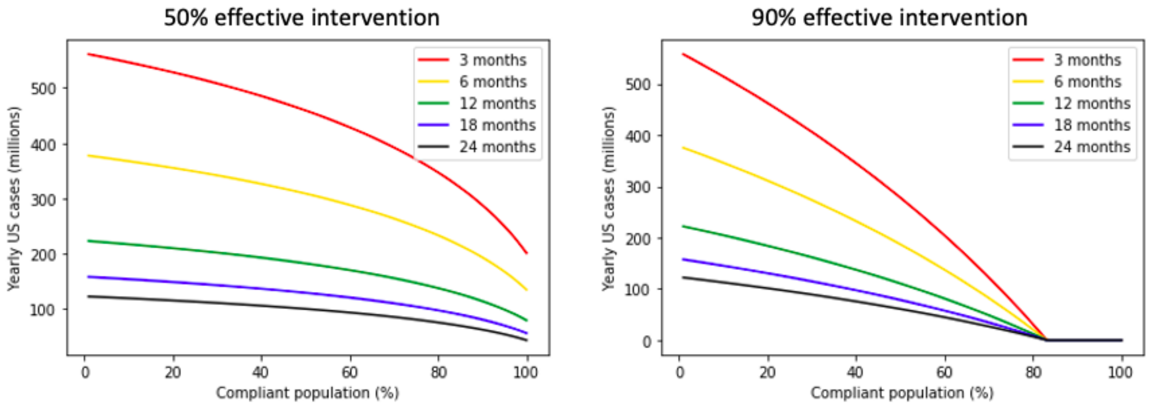


**Figure S3:** Duration of natural immunity impacts yearly disease burden but not the compliance threshold for complete suppression. Yearly predicted US cases at steady-state under a range of assumptions about the duration of natural immunity.

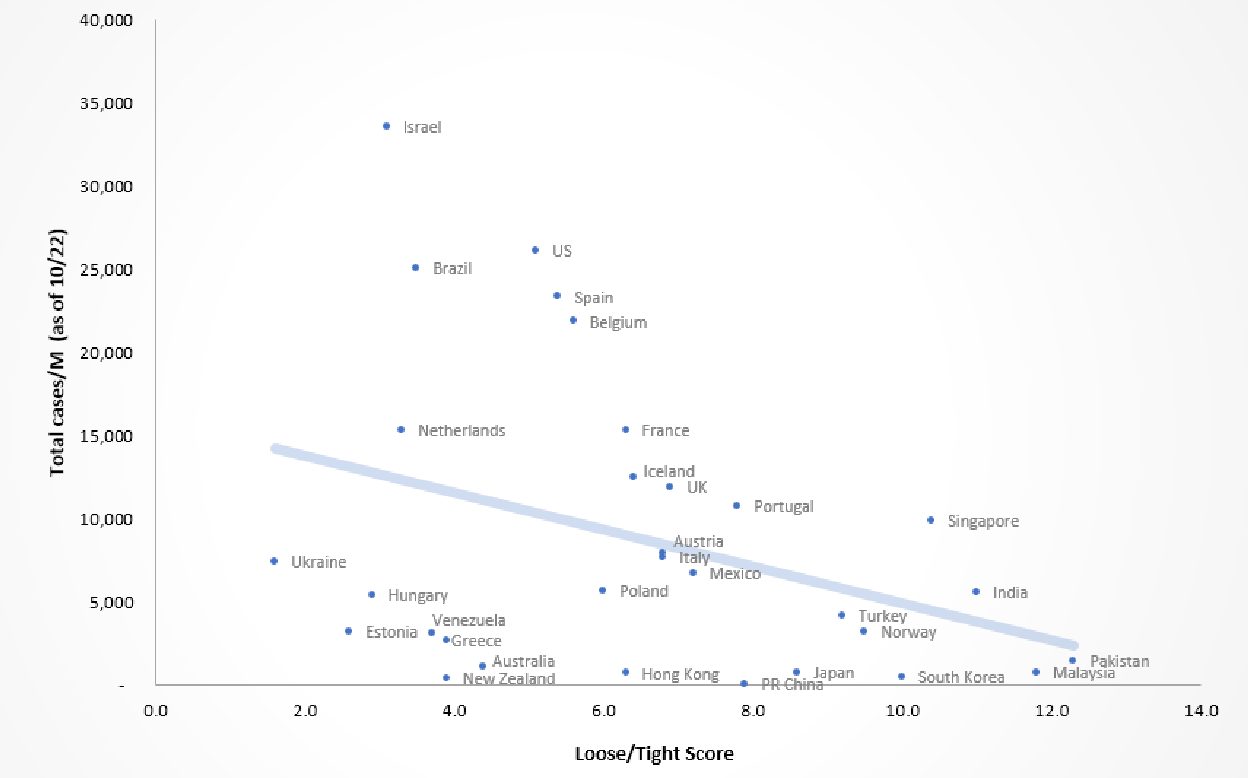


| | |
|---|---|
| Pearsons R | 0.36 |
| p-value | 0.042983 |

**Figure S4:** Association between loose-tight score and COVID-19 cases per capita (*7*). There is a weak association with loose-tight cultures and total case count observed based on reported case counts as of October.

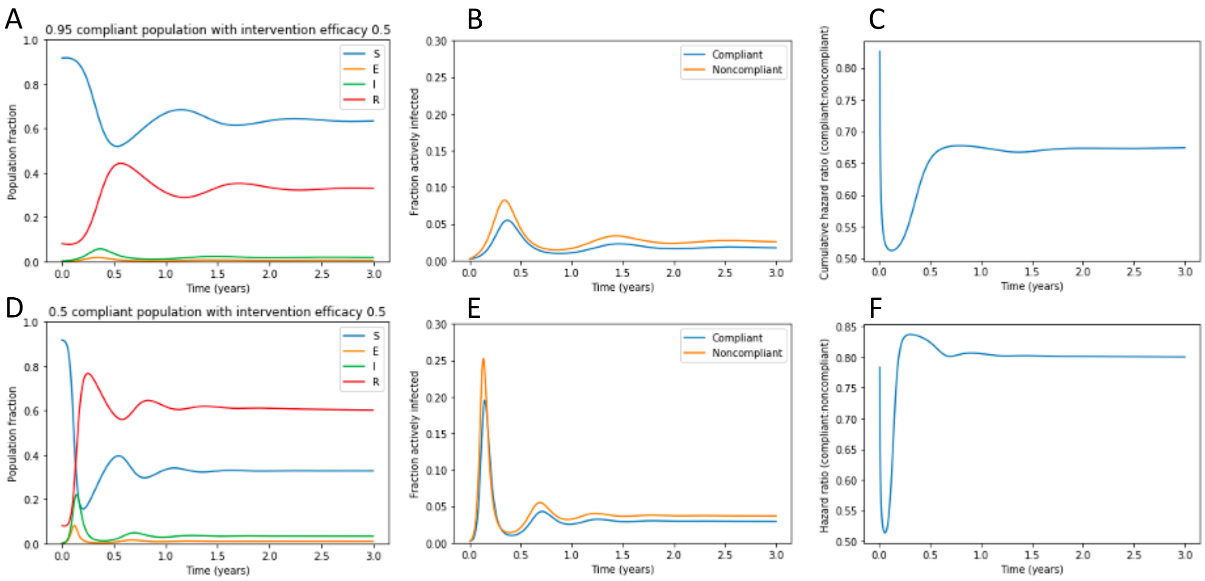


**Figure S5.** Complement to Figure 2, with simulated 6-month duration of natural immunity. Panels A and D represent the fraction of the population, including both compliant and noncompliant individuals, that is susceptible, exposed, infectious, and recovered populations over time after a return to pre-pandemic conditions under (A-C) 95% compliance or (D-F) 50% compliance with a 50% effective intervention. Panels B and E demonstrate the fraction of compliant and noncompliant individuals who are infected over time. Panels C and F demonstrate the cumulative hazard ratio for infection in noncompliant versus compliant individuals.

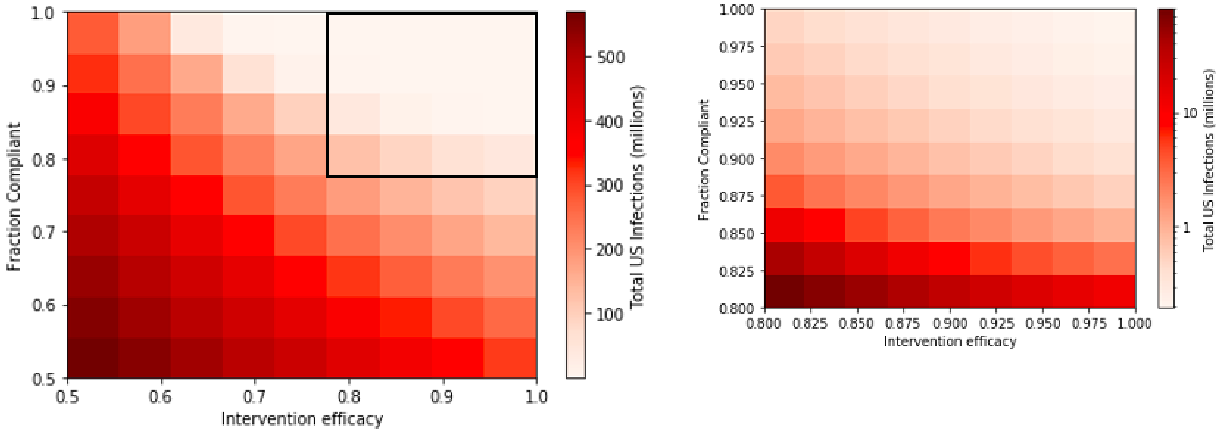


**Figure S6.** Complement to Figure 3, with simulated 6-month duration of natural immunity. Total US COVID-19 infections in the next year under interventions with varying efficacy and compliance. Black box on panel A shows region expanded in panel B. Panel B is displayed on a log scale.

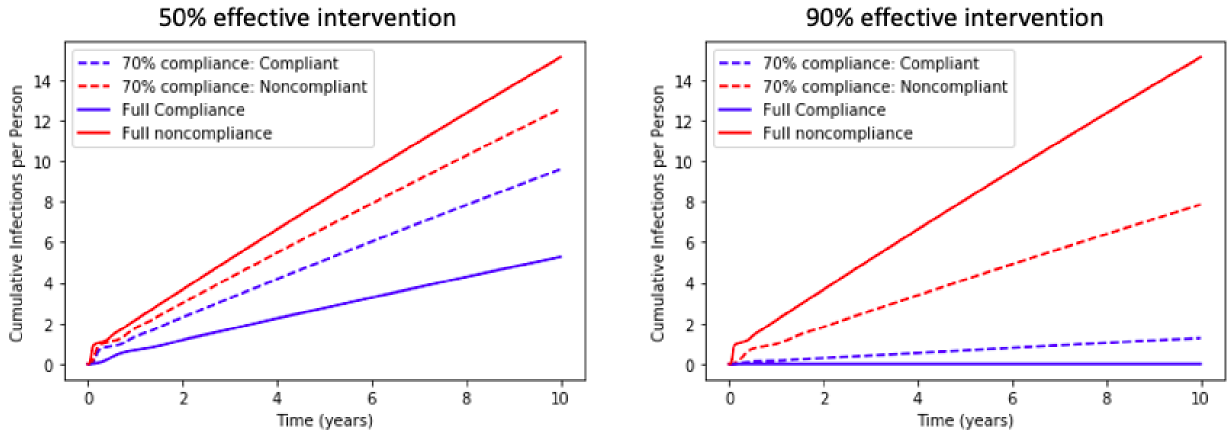


**Figure S7.** Complement to Figure 4, with simulated 6-month duration of natural immunity. Cumulative infections per individual under a 50% or 90% effective intervention. Three scenarios are simulated: full noncompliance, full compliance, and 70% compliance (with outcomes for compliant and noncompliant individuals shown).

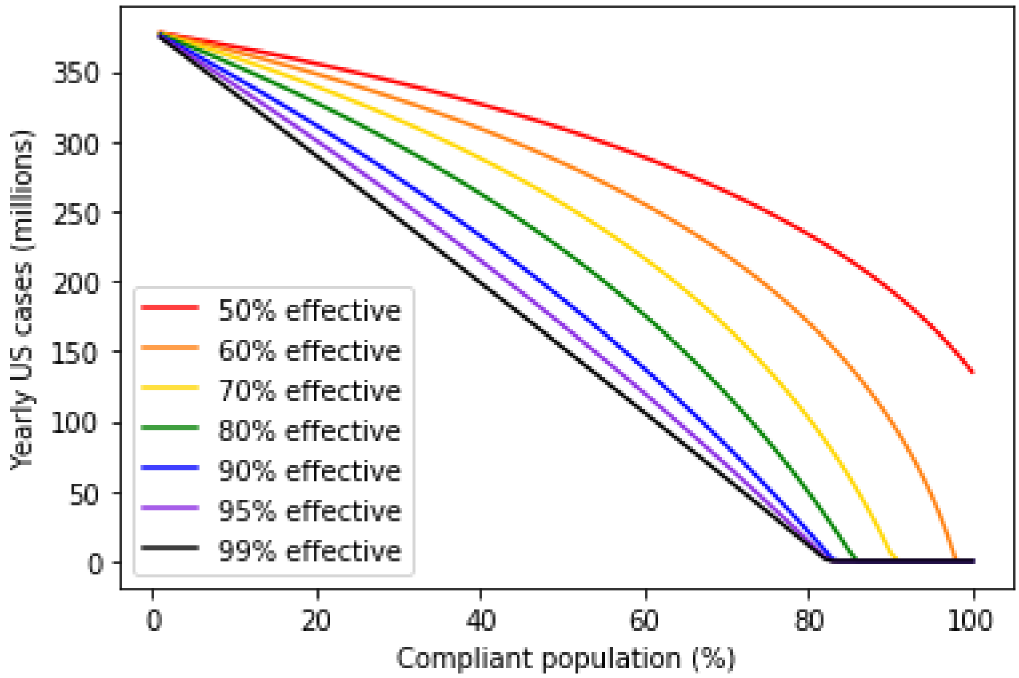


**Figure S8.** Complement to Figure 5, with simulated 6-month duration of natural immunity. Yearly US cases at steady-state under interventions with varying degrees of efficacy and compliance.

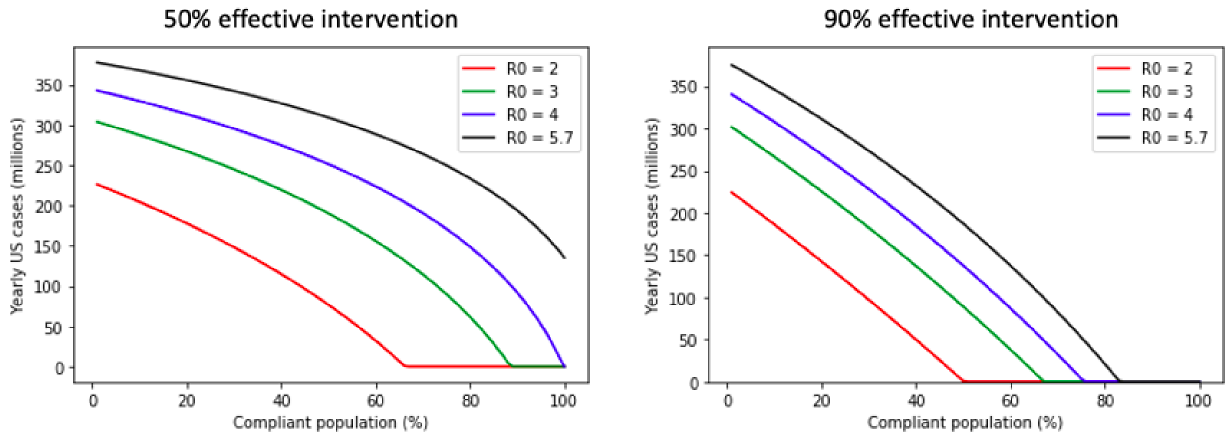


**Figure S9.** Complement to Figure S2, with simulated 6-month duration of natural immunity. Yearly US cases as a function of compliance for hypothetical scenarios in which $R_0$ = 2, 3, 4 or 5.7.

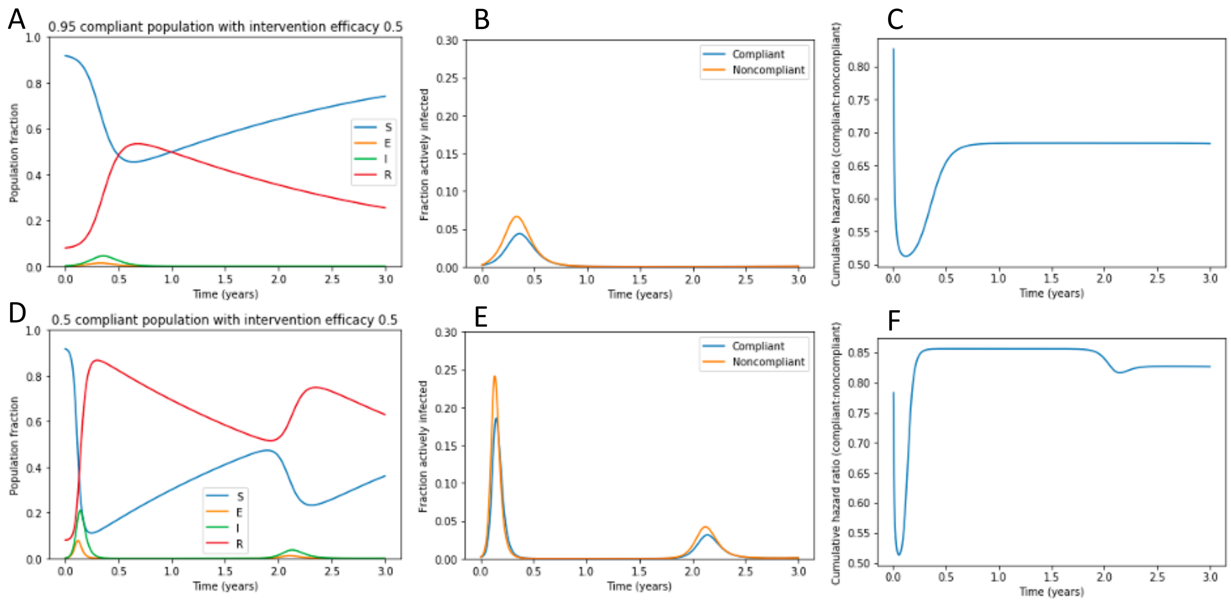


**Figure S10.** Complement to Figure 2, with simulated 36-month duration of natural immunity. Panels A and D represent the fraction of the population, including both compliant and noncompliant individuals, that is susceptible, exposed, infectious, and recovered populations over time after a return to pre-pandemic conditions under (A-C) 95% compliance or (D-F) 50% compliance with a 50% effective intervention. Panels B and E demonstrate the fraction of compliant and noncompliant individuals who are infected over time. Panels C and F demonstrate the cumulative hazard ratio for infection in noncompliant versus compliant individuals.

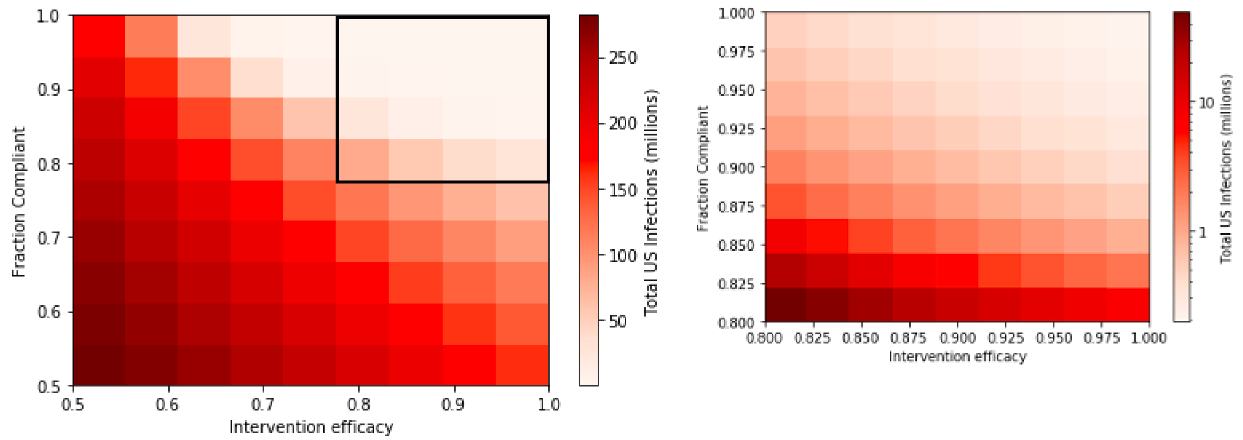


**Figure S11.** Complement to Figure 3, with simulated 36-month duration of natural immunity. Total US COVID-19 infections in the next year under interventions with varying efficacy and compliance. Black box on panel A shows region expanded in panel B. Panel B is displayed on a log scale.

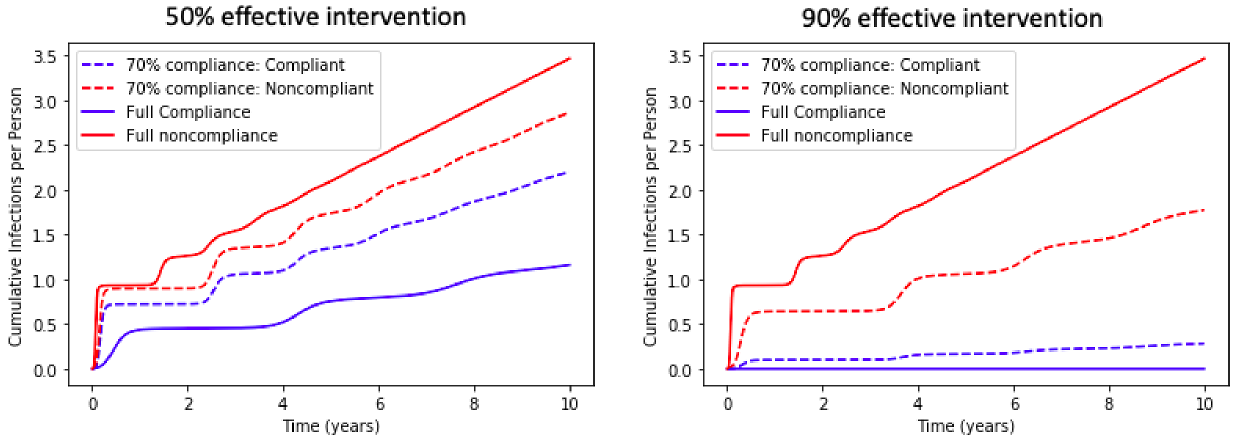


**Figure S12.** Complement to Figure 4, with simulated 36-month duration of natural immunity. Cumulative infections per individual under a 50% or 90% effective intervention. Three scenarios are simulated: full noncompliance, full compliance, and 70% compliance (with outcomes for compliant and noncompliant individuals shown).

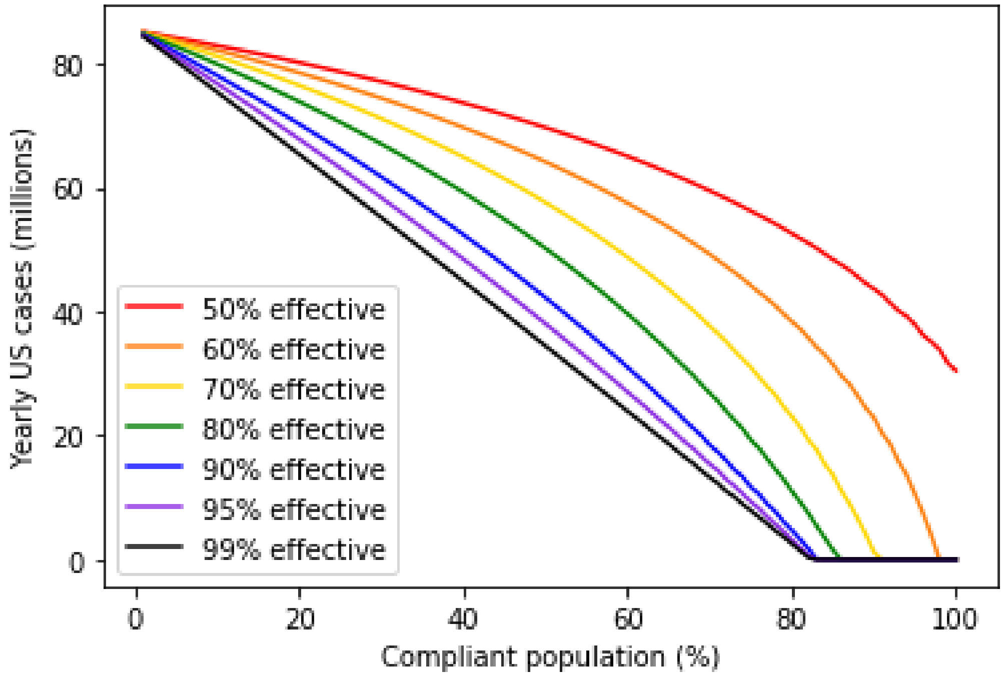


**Figure S13.** Complement to Figure 5, with simulated 36-month duration of natural immunity. Yearly US cases at steady-state under interventions with varying degrees of efficacy and compliance.

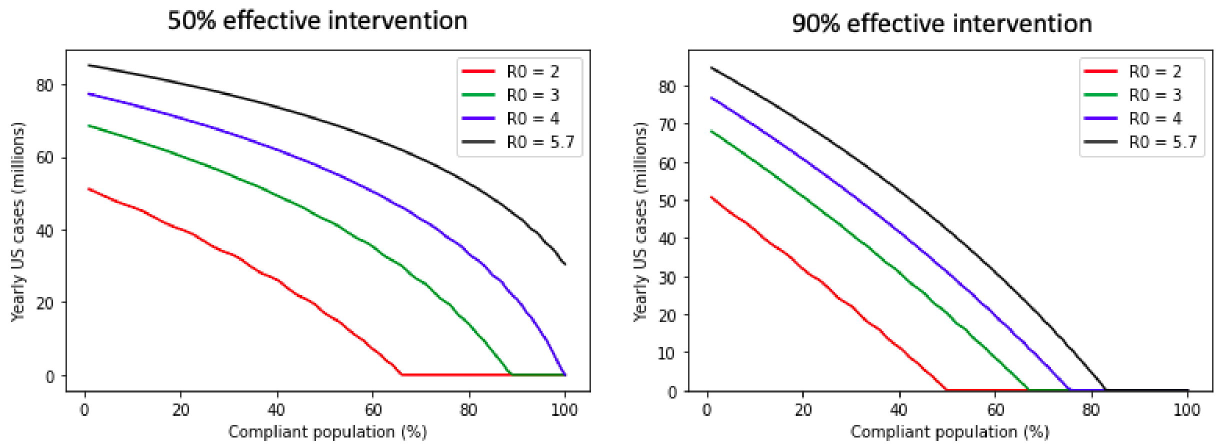


**Figure S14.** Complement to Figure S2, with simulated 36-month duration of natural immunity. Yearly US cases as a function of compliance for hypothetical scenarios in which $R_0$ = 2, 3, 4 or 5.7.

### References


1. South Dakota Coronavirus: 76,142 Cases and 849 Deaths (COVID-19 ) - Worldometer, (available at https://www.worldometers.info/coronavirus/usa/south-dakota/).

2. South Dakota Population 2020 (Demographics, Maps, Graphs), (available at https://worldpopulationreview.com/states/south-dakota-population).

3. Epidemiology and transmission dynamics of COVID-19 in two Indian states | Science, (available at https://science.sciencemag.org/content/370/6517/691).

4. D. K. Chu, E. A. Akl, S. Duda, K. Solo, S. Yaacoub, H. J. Schünemann, Physical distancing, face masks, and eye protection to prevent person-to-person transmission of SARS-CoV-2 and COVID-19: a systematic review and meta-analysis. *Lancet*. **395**, 1973–1987 (2020).

5. Assessing the Age Specificity of Infection Fatality Rates for COVID-19: Systematic Review, Meta-Analysis, and Public Policy Implications | medRxiv, (available at https://www.medrxiv.org/content/10.1101/2020.07.23.20160895v7).

6. NHTSA, 2018 Data: Alcohol-Impaired Driving, (available at https://crashstats.nhtsa.dot.gov/Api/Public/ViewPublication/812864).

7. M. J. Gelfand, J. L. Raver, L. Nishii, L. M. Leslie, J. Lun, B. C. Lim, L. Duan, A. Almaliach, S. Ang, J. Arnadottir, Z. Aycan, K. Boehnke, P. Boski, R. Cabecinhas, D. Chan, J. Chhokar, A. D'Amato, M. Ferrer, I. C. Fischlmayr, R. Fischer, M. Fülöp, J. Georgas, E. S. Kashima, Y. Kashima, K. Kim, A. Lempereur, P. Marquez, R. Othman, B. Overlaet, P. Panagiotopoulou, K. Peltzer, L. R. Perez-Florizno, L. Ponomarenko, A. Realo, V. Schei, M. Schmitt, P. B. Smith, N. Soomro, E. Szabo, N. Taveesin, M. Toyama, E. Van de Vliert, N. Vohra, C. Ward, S. Yamaguchi, Differences between tight and loose cultures: a 33-nation study. *Science*. **332**, 1100–1104 (2011).
